# Synthetic Population Generation with Public Health Characteristics for Spatial Agent-Based Models

**DOI:** 10.1101/2024.09.18.24312662

**Authors:** Emma Von Hoene, Amira Roess, Hamdi Kavak, Taylor Anderson

## Abstract

Agent-based models (ABMs) simulate the behaviors, interactions, and disease transmission between individual “agents” within their environment, enabling the investigation of the underlying processes driving disease dynamics and how these processes may be influenced by policy interventions. Despite the critical role that characteristics such as health attitudes and vaccination status play in disease outcomes, the initialization of agent populations with these variables is often oversimplified, overlooking statistical relationships between attitudes and other characteristics or lacking spatial heterogeneity. Leveraging population synthesis methods to create populations with realistic health attitudes and protective behaviors for spatial ABMs has yet to be fully explored. Therefore, this study introduces a novel application for generating synthetic populations with protective behaviors and associated attitudes using public health surveys instead of traditional individual-level survey datasets from the census. We test our approach using two different public health surveys (one national and the other representative of the study area, Virginia, U.S.) to create two synthetic populations representing individuals aged 18 and over in Virginia, U.S., and their COVID-19 vaccine attitudes and uptake as of December 2021. Results show that integrating public health surveys into synthetic population generation processes preserves the statistical relationships between vaccine uptake and attitudes in different demographic groups while capturing spatial heterogeneity at fine scales. This approach can support disease simulations that aim to explore how real populations might respond to interventions and how these responses may lead to demographic or geographic health disparities. Our study also demonstrates the potential for initializing agents with variables relevant to public health domains that extend beyond infectious diseases, ultimately advancing data-driven ABMs for geographically targeted decision-making.

**Author Summary:** In this study, we introduce a new method for generating synthetic populations of individuals or “agents” with characteristics that include health protective behaviors and attitudes, which are crucial for modeling disease spread. Traditional methods for parameterizing agents often overlook the complex relationships between demographic factors and health behaviors like vaccination. Additionally, detailed spatial data capturing these behaviors are limited, meaning agent behaviors are more uniform across geographic space. By fitting public health surveys with spatially aggregated census data, we created more realistic agent populations for disease spread simulations. We focused on Virginia, U.S. and generated a population with COVID-19 vaccine uptake and attitudes as of December 2021. Our results show that this approach captures the statistical relationships between demographic variables and vaccine uptake, along with the spatial variation in these behaviors. We also show that using national survey data is comparable to using local survey data representative of Virginia collected in 2021. The approach is flexible so that it can be applied to various public health studies beyond just infectious diseases. Our work highlights the potential of public health surveys for enhancing synthetic population generation, offering a valuable approach for initializing models with more realistic populations to explore public health challenges.

## 1. Introduction

Agent-based models (ABMs) are commonly used to simulate the spread of infectious diseases caused by viruses, including COVID-19 virus (1–4), influenza (5,6), and the chickenpox virus (7,8). Unlike traditional epidemiological models, such as the Susceptible-Infectious-Recovered (SIR) model and its variants, ABMs use a bottom-up approach that simulates the behaviors, interactions, and subsequent transmission of disease between individual “agents” within their environment (9,10). This approach allows for the investigation of the underlying processes driving disease dynamics and how these processes may be influenced by policy interventions (11).

Given the important role of demographic characteristics such as age and income (12), household structures (13–15), activity patterns and co-location (16) in disease dynamics, most ABMs of disease spread attempt to incorporate these attributes when initializing agents. For example, children often participate in activities like attending school or recreational events, where they interact with many other individuals and are more likely to contract pathogens that can then be transmitted to parents or grandparents living in the same household (17). While some studies use random functions or fixed values to assign agent attributes, population synthesis approaches can utilize spatially aggregated census data and individual-level survey data (18) from sources like household travel surveys (19) or census microdata (20) to create a complete agent population with relevant attributes, including household structures, thus accurately capturing these transmission pathways within the model.

Disease dynamics are also shaped by the uptake of protective behaviors by the population, such as wearing masks, getting vaccinated, and staying home when sick, which can reduce the likelihood of negative health outcomes (21). In addition to social norms and physical or financial barriers, an individual’s attitudes, beliefs, and perceptions significantly affect their decision to engage in protective behaviors. Although traditionally overlooked in ABMs of disease spread (22–24), the COVID-19 pandemic spurred on a widespread effort to better represent health behaviors and their dynamics into epidemiological models (25–28). This paper argues that a synthetic population generation approach capable of initializing agent populations with a realistic set of attitudes and protective behaviors can support such ABMs that aim to simulate behavior dynamics influenced by these attributes.

A typical approach in current ABMs is that protective behaviors or related attitudes are assigned to agents with some probability, either based on a hypothetical scenario or based on aggregate data measuring the real characteristics of the population. For example, Rafferty et al. (7) use an ABM to simulate the impact of dose timing, coverage, and waning of immunity on chickenpox disease outcomes in Alberta, Canada. They initialize the population with vaccination attitudes based on aggregate data (65% acceptance, 30% hesitant, 5% reject). However, this approach ignores the statistical relationships between vaccine attitudes and other individual demographic, cultural, or political characteristics, that synthetic population generation approaches aim to preserve.

In another example, Pandey et al. (29) use an ABM to examine the effect of bivalent boosters on COVID-19 outcomes, assuming a coverage of 59%, 51%, 38%, 54%, and 75% for age groups 5-11, 12- 17, 18-49, 50-64 and 65+, respectively, informed by historical influenza data. While their model more accurately captures the relationship between booster coverage and age, whereby age 65+ are more likely to accept a booster, the study assumes spatially uniform uptake across New York City. This assumption of uniformity is common, especially since health data are often not available at granularities finer than county or state, meaning that spatial heterogeneities can only be captured at these levels. While numerous ABMs have been developed to simulate the adoption of protective behaviors or the spread of beliefs, attitudes, perceptions towards vaccines over space and time, the use of synthetic population approaches to initialize an agent population with these characteristics has yet to be explored.

Therefore, the purpose of this study is to investigate how synthetic population generation approaches can be expanded to create agent populations with attitudes and initial adoption of protective behaviors, along with their spatial distributions. Specifically, we aim to replace datasets commonly used in synthetic population generation that provide individual-level data from samples with coarser geographic resolution, such as the Census Bureau’s Public Use Microdata Sample (PUMS) (20), with public health surveys. Using COVID-19 as a case study, we explore the potential for this approach by generating a synthetic population representing Virginia, U.S. and their vaccine attitudes and uptake as of December 2021. We obtain real vaccine uptake for Virginia at the CT level for the same point in time to validate our results, comparing the populations generated by two different public health surveys: one national, and the other representative of Virginia.

## 2. Background

With the growing use of ABMs across disciplines such as economics, geography and biology (30), a wealth of synthetic population generation methods have been developed to create agent populations. These populations serve as simplified microscopic representations of the targeted population, reflecting individuals and their socio-demographic characteristics relevant to the study (31). The emergence of synthetic population generation approaches is largely due to several factors, including privacy restrictions that prevent access to detailed individual-level data at fine spatial scales, the ability of ABMs to simulate social dynamics and behaviors which are connected with individual attributes, and advancements that have made ABMs more data-informed and effective as predictive tools for decision support (32,33).

Synthetic population generation methods vary in complexity and are broadly categorized into two main approaches: Combinatorial Optimization (CO) and Synthetic Reconstruction (SR). CO focuses on replicating real entities by reweighting an existing dataset to match individual profiles. In contrast, SR, which is more commonly used and well-established, generates populations through random sampling from known distributions of demographic characteristics or estimated joint distributions using deterministic re-weighting algorithms like Iterative Proportional Fitting (IPF) (31,34). Given the extensive literature on population synthesis, we provide only a brief background to support understanding of our proposed method. For a comprehensive review of population generation approaches for ABMs, see Chapuis et al. (31).

IPF is the most widely used approach for generating synthetic populations due to its long-standing presence and reliability in literature, computational efficiency, and its methodological simplicity (35). The algorithm adjusts each cell in an n-dimensional matrix, which represents the distribution of attributes, based on known marginal controls. It starts with sample data to initialize the matrix and then iteratively updates the cells to match the specified contingency dimensions (36). Originally introduced by Deming and Stephan (37) to adjust contingency tables to fit with known marginal distributions, IPF has been extensively refined by researchers to improve its application for population synthesis. For instance, Beckman et al. (38) first established the methods for using IPF with PUMS data, where joint distributions of household attributes were derived by integrating sample frequency tables from PUMS data with marginal distributions from Census Summary Files, and then randomly selecting households based on these estimates to create a synthetic population.

Synthetic population generation approaches, such as those using IPF to initialize an agent population within a spatial ABM, typically combine spatially aggregate and disaggregated individual-level data to statistically match both the joint distributions found in the individual-level data with the marginal totals in the spatially aggregate data (18). Spatially aggregate data captures marginal totals of populations across a set of categories such as gender, age, and race within different geographic zones (e.g. census tracts, dissemination areas) in a study area. This data allows for analysis of populations and their spatial distributions with relatively fine granularity while preserving privacy by presenting only marginal totals (e.g., total population aged 65+, or total population that is white) rather than joint distributions across multiple attributes (e.g., total population aged 65+ and white). Disaggregated individual-level survey data contains samples of anonymized records of real individuals and their demographic characteristics. Although this data captures the joint distributions among individual attributes, it represents only a small sample from a large geographic area (e.g., a state or the entire country), which protects individual identities and prevents inference of the spatial distribution of the sample population. Examples of such datasets include PUMS in the U.S., and similar datasets available in other countries, such as Public Use Microdata Files (PUMFs) in Canada.

Synthetic population generation, particularly SR approaches, involves both fitting and allocation. During the fitting stage, IPF is used to align individual-level sociodemographic data with spatially aggregated constraints, generating fractional weights for entities such as households or individuals in each geographic zone. Because IPF outputs fractional weights, allocation is required to produce a discrete set of agent counts that replicates individuals (32). The fractional weights are converted into integer weights through a process known as ‘integerisation’, which can be performed using various approaches such as simple rounding, thresholding, proportional probabilities, or truncate, replicate, sample (TRS) (see Lovelace and Ballas (39) for a review on these methods). ‘Integerisation’ is followed by expansion, where each individual is represented as a record with a geographic zone, and the matching attributes for that individual from the original survey dataset is carried over (39).

IPF has been used to create agent populations in various spatial ABMs, such as those for disaster management and recovery (40), though it is most commonly used in urban and transportation modeling. However, in the context of spatial ABMs for infectious disease spread, there are few dedicated population synthesis methods or studies utilizing well-established techniques such as IPF (41). This is likely because ABMs take significant time to develop and are often designed for specific objectives, such as understanding the impact of policy guidelines and health behaviors on infectious disease dynamics (42–44), proposing general or behavioral frameworks for epidemiological models (45,46), or forecasting disease transmission (48,49). This gap is particularly significant as these models are valuable for informing policy, yet generating populations with detailed individual characteristics and health behaviors often remains overlooked, despite their critical role in influencing disease transmission. To our knowledge, no specific synthetic population generation method for spatial ABMs of disease spread has yet been developed to capture both individual attitudes and initial adoption of protective behaviors, along with the spatial heterogeneities in these characteristics. Therefore, there is a need for a flexible synthetic population generation approach that accurately initializes agent populations with attitudes, beliefs, perceptions and initial adoption of protective behaviors, as well their spatial distributions. By proposing a targeted population synthesis method that derives these individual attributes from public health surveys, this approach can be adapted for various public health applications, including infectious diseases, smoking, and other health challenges, across different scales and locations.

## 3. Materials and Methods

Our approach is presented in Figure 1. First, individual-level survey data and spatially aggregate data are used as input data for the population synthesis of agents with demographic characteristics. Our approach extends traditional synthetic population generation approaches by allowing for vaccination status and attitudes to be carried over at the replication stage. We compare our approach with a null model, which uniformly assigns vaccine uptake likelihood based on county level vaccine uptake data. Our validation involves comparing the vaccination rates in the synthetic population to those in the real population for each census tract. This comparison is conducted for populations generated using two different public health surveys (a local survey and a national survey) and their respective null models. The data and the methods are described in detail in the following sections. The code written in the R scripting language and the data for the synthetic population generation approach and the validation is available at the GitHub repository https://github.com/evonhoene/Population-Generation-for-Public-Health-ABMs.

**Figure 1.**
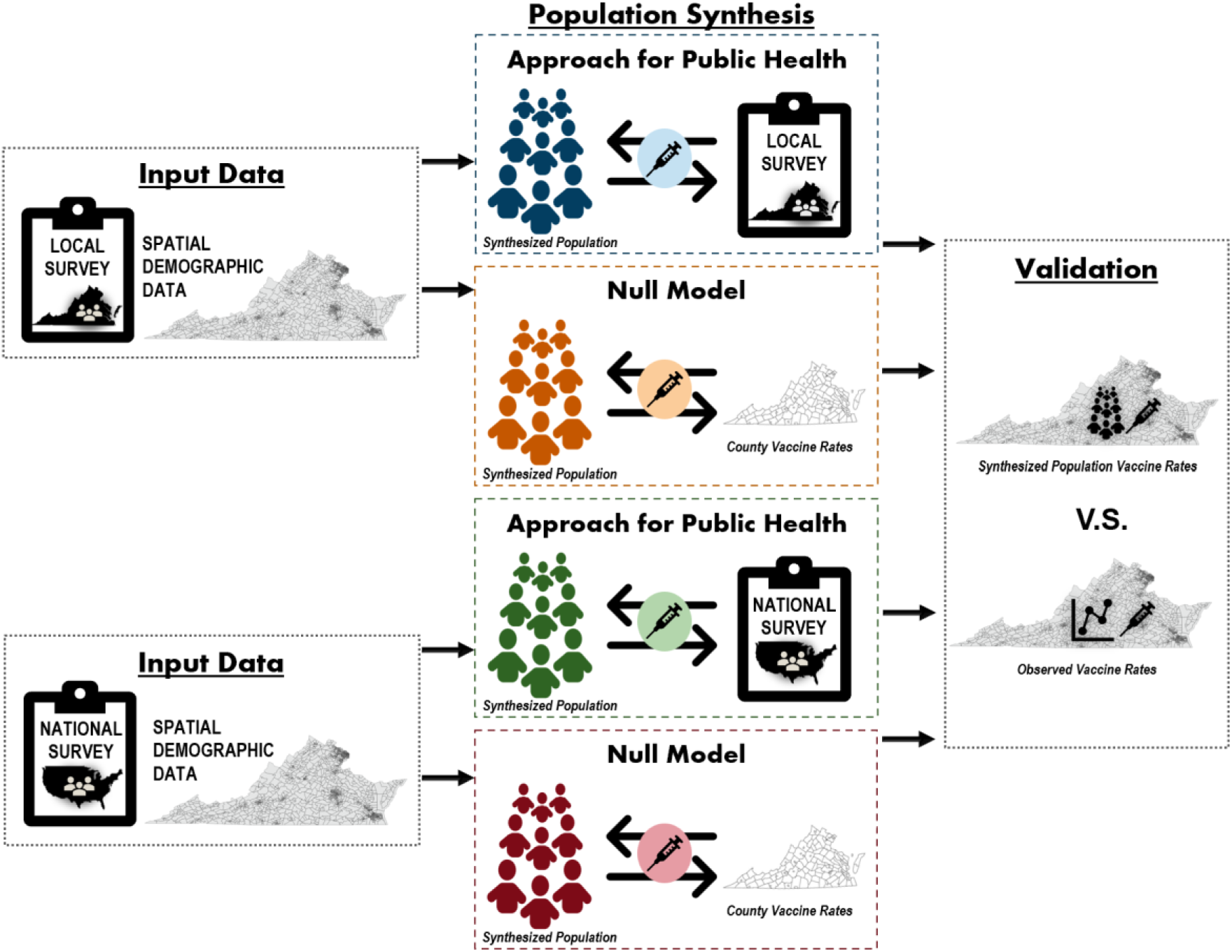
Overview of the approach used in the study.

### 3.1. Input Data

Given that IPF is a well-established, efficient, and straightforward method for synthetic population generation, we use it to ensure the flexibility of our proposed approach for various study applications. This method requires both spatially aggregated demographic data and individual-level survey data. For the spatially aggregated data, we use census tract (CT) data from the Census Bureau’s American Community Survey (ACS) (50) that captures marginal totals for sociodemographic variables. We focus on gender, race, age, education, and income variables for individuals aged 18 and over, as these factors significantly influence COVID-19 vaccine uptake (51). While the ACS provides marginal totals for individuals across different categories of gender, race, age, and education, income data is reported as the percentage of households within each CT that fall into specific income brackets. To estimate individual income levels, we calculate the proportion of the total population aged 18+ that would fall into each income bracket, assuming a household size of 1. We use 2021 data specifically for Virginia CTs and exclude records with missing or zero values for any variable, resulting in a final dataset of N = 2162. Descriptive statistics for the variables collected from the ACS dataset are presented in Table 1.

**Table 1.** Descriptive statistics summarizing the demographic distribution within Virginia Census Tracts.

| Variable: Descriptor | Mean % | Minimum % | Median % | Maximum % |
| --- | --- | --- | --- | --- |
| Gender: Male | 48.70285 | 0 | 48.73321 | 100 |
| Gender: Female | 51.29715 | 0 | 51.26679 | 100 |
| Race/Ethnicity: White | 63.14219 | 0 | 66.38809 | 100 |
| Race/Ethnicity: Black | 19.90094 | 0 | 12.77431 | 100 |
| Race/Ethnicity: Hispanic | 7.914201 | 0 | 5.032508 | 80.2409 |
| Race/Ethnicity: Other | 9.065319 | 0 | 5.853839 | 62.07447 |
| Age: 18-29 | 20.41858 | 0 | 17.94349 | 98.78631 |
| Age: 30-49 | 33.61571 | 0 | 33.13478 | 71.48159 |
| Age: 50-64 | 25.43375 | 0 | 25.9862 | 100 |
| Age: 65 and over | 20.53196 | 0 | 19.91834 | 100 |
| Education: Bachelor's degree or higher | 17.29956 | 0 | 14.93085 | 53.30806 |
| Education: No Bachelor's degree | 82.70044 | 46.69194 | 85.06915 | 100 |
| Income: Less than \$25,000 | 14.95912 | 0 | 11.69449 | 100 |
| Income: \$25,000 - \$49,999 | 17.3773 | 0 | 17.09721 | 63.68978 |
| Income: \$50,000 - \$99,999 | 28.74916 | 0 | 28.9908 | 100 |
| Income: Greater than \$100,000 | 39.18442 | 0 | 35.24403 | 100 |

Our approach replaces the traditional individual-level samples captured by censuses used in synthetic population generation (e.g. the PUMS in the US) with public health surveys. We compare the results of our approach using two surveys, one that is one that is representative of Virginia and one that is national and publicly available, as follows:

1. *Local Survey*: This survey, collected by researchers is representative of the Commonwealth of Virginia and includes data on demographics as well as beliefs, attitudes, and perceptions related to COVID-19 and protective behaviors. The sample was recruited by Climate Nexus Polling (August 15-31, 2021), using several market research panels. Participants were recruited using stratified sampling methods. Compensation for participants depended on the specific market research panel and respondents’ preferences (e.g., cash, gift cards, reward points). Sampling weights accounted for small deviations from the pre-selected census parameters. The dataset includes N = 3,528 respondents. The descriptive statistics for the data are provided in Table 2. De-identitied data are available upon request. This project to collect the local survey data was considered exempt by the George Mason University IRB (IRB 1684418-3).
2. *Household Pulse Survey (HPS)*: This publicly available national survey, obtained from the US Census Bureau (52), measures the impact of emergent social and economic issues on households across the country, including COVID-19 vaccinations. The HPS also collects data on core demographic characteristics from respondents aged 18 and older. We use data from HPS Week 41, covering December 29, 2021, to January 10, 2022. Records missing data for one or more variables were removed (e.g., vaccine decision, household income), resulting in a total of N = 63,180 respondents. Given the large size of the HPS dataset, we use stratified sampling to reduce the sample to 3,500 to match the size of the *local* survey. As described in Table 2, the HPS data shows a bias, with 91.19% of respondents reporting being vaccinated. At the same time, publicly available county-level vaccination data from the CDC (53) indicates that only 50.2% of Virginians were vaccinated by December 30, 2021. To correct this bias in our stratified sample of 3,500 records, we adjust so that 50% of the sample is vaccinated while preserving the representation of all other variables.

**Table 2.** Comparative distribution of respondent characteristics from the individual-level surveys.

| Variable: Descriptor | % Respondents from HPS | % Respondents from Local Survey |
| --- | --- | --- |
| Gender: Male | 41.28 | 44.76 |
| Gender: Female | 58.72 | 55.24 |
| Race/Ethnicity: White | 75.06 | 74.12 |
| Race/Ethnicity: Black | 7.01 | 10.15 |
| Race/Ethnicity: Hispanic | 9.16 | 10.86 |
| Race/Ethnicity: Other | 8.78 | 4.88 |
| Age: 18-29 | 9.15 | 16.10 |
| Age: 30-49 | 36.93 | 39.12 |
| Age: 50-64 | 29.15 | 20.29 |
| Age: 65 and over | 24.76 | 24.49 |
| Education: Bachelor's degree or higher | 41.86 | 36.96 |
| Education: No Bachelor's degree | 58.14 | 63.04 |
| Income: Less than \$25,000 | 11.74 | 52.15 |
| Income: \$25,000 - \$49,999 | 19.72 | |
| Income: \$50,000 - \$99,999 | 31.03 | 28.66 |
| Income: Greater than \$100,000 | 37.51 | 19.19 |
| Vaccination: Yes | 91.19 | 65.33 |
| Vaccination: No | 8.81 | 34.67 |

Each survey is used to generate a separate set of synthetic population. In the surveys, while some of the individual-level data is measured on a continuous scale (e.g. age), other data are measured categorically, which results in varying levels of detail between the two synthetic populations, depending on the questions asked. For example, when asking about income, the local survey allows respondents to select <$50,000, $50,000-$99,999, and >$100,000. On the other hand, the HPS allows respondents to select <$25,000 and $25,000-$49,999, $50,000-$99,999, and >$100,000, allowing for slightly more detailed agent characteristics related to income. In any case, to be used in the IPF process, the data measured by the individual-level survey must be able to be fall under the categories in the spatially aggregated ACS data. This was possible for attributes including gender, race and ethnicity, age, education, and income. Descriptive statistics for the variables captured by the individual-level surveys and their categories from the two survey datasets are outlined in Table 2.

Our validation approach uses CT data capturing marginal totals for real vaccine uptake aged 12+ in Virginia as of December 30, 2021. This data is not publicly available and was acquired by request from the Virginia Department of Public Health. The department has since scaled down its operations, and this data is no longer accessible, even upon request. The vaccine uptake data is available for 1,601 CTs for which we generate a population. Furthermore, 9 records indicated that vaccine uptake was greater than 100% and were removed. As such our validation only focuses on CTs where data is available, and vaccine uptake is less than or equal to 100% (N=1592).

### 3.2. Population Synthesis

#### Synthetic population generation approach for public health

We use IPF to generate approximately 6 million agents representing the population of Virginia aged 18 and over. This includes generating one population based on the HPS and another using the local survey. Our approach is detailed in Figure 2. IPF computes a weight for every individual in the survey based on how well their characteristics represents the *age, gender, race, income, and education* distributions found in the CT population. These weights are then processed using the TRS ‘integerisation’ method (39), which involves truncating all weights to integers and using these as the counts of each individual type in the zone, followed by sampling to achieve the correct population size based on the probabilities corresponding to the decimal weights. Simply, this approach converts the weights to integers that describe how many times that individual respondent in the survey should be replicated as an agent in the CT. This process is repeated for each CT in the study area. Following this, expansion is conducted to create the final dataset, where each record corresponds to an individual and their CT. By replacing the PUMS with the public health surveys, the demographic variables and every other variable captured by the surveys including a COVID-19 vaccination status as well as attitudes, perceptions, and beliefs related to vaccine are carried over in the sampling and replication stage.

**Figure 2.**
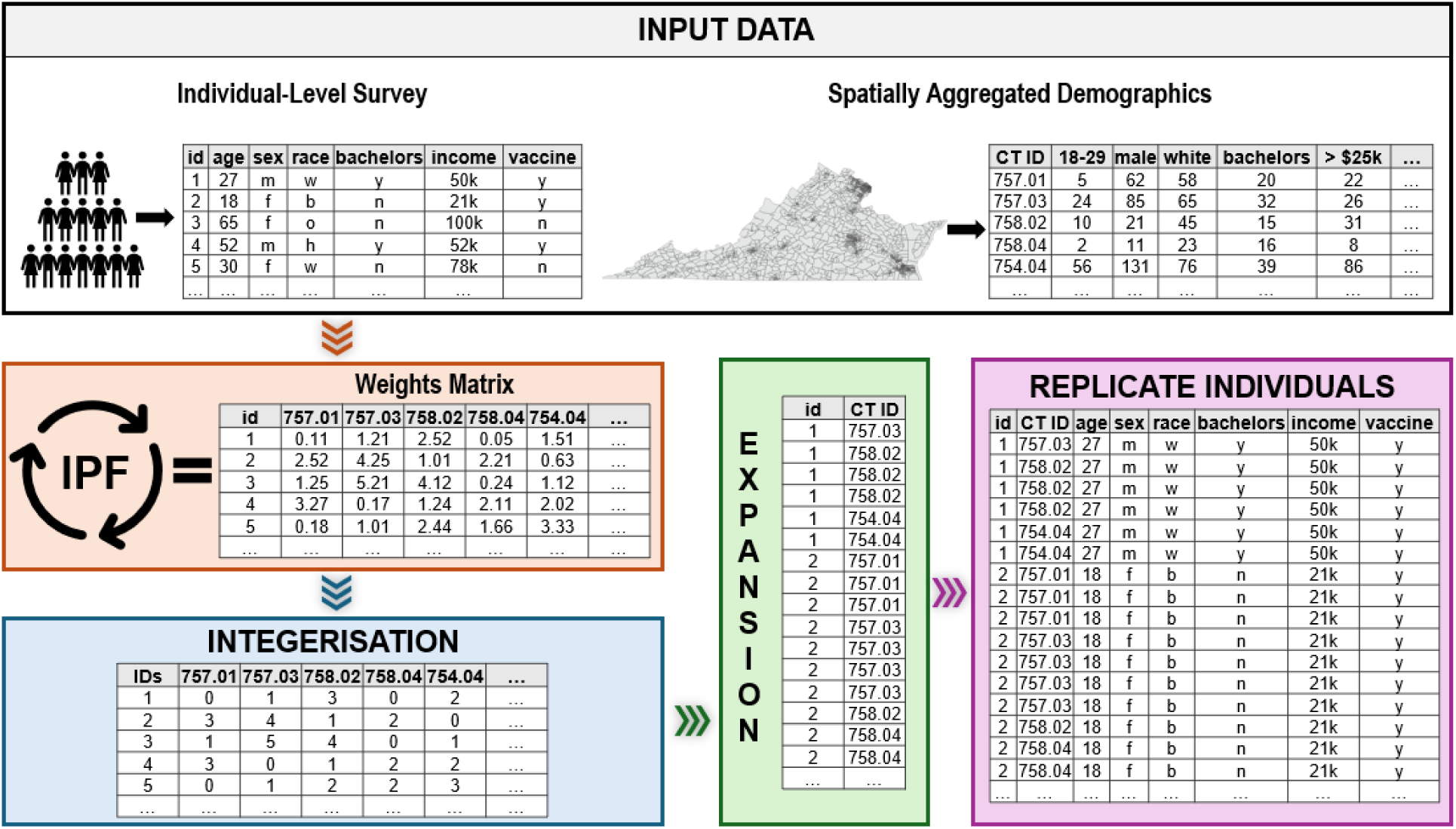
Overview of the synthetic population generation approach for public health.

#### Null model

We compare the results of our public health synthetic population generation approach with a null model that serves as a baseline. Two distinct populations were generated using the null model, corresponding to the HPS and local survey datasets. With the null model, the IPF method fits the individual level demographic data from the surveys with the CT data, creating a population of agents with age, gender, race, income, and education characteristics for each CT in Virginia. However, since vaccine uptake information is only publicly available at county-level, the null model uses this data to impose vaccination uniformly on agents in the same county. For example, as of December 30, 2021, 84.5% of individuals aged 18+ living in Fairfax County were vaccinated (53). Therefore, all agents generated for Fairfax County in the null model are assigned a vaccination likelihood of 84.5%. This is a common approach in ABM to initialize agents with health variables such as vaccine uptake.

#### Validation

We compare the spatial and statistical patterns of the simulated vaccine uptake with the observed vaccine uptake percentage at the CT level and with the individual level survey data for the same time period. Although the population is generated for all CTs in the study area, validation is only possible for CTs where real vaccine uptake data is available and where vaccine uptake is less than or equal to 100%.

## 4. Results

We evaluate the observed and simulated percentages of gender, race, age, education, income, and vaccine status variables for Virginia CTs (N = 1,592) in the populations generated using the HPS and the local survey, using the following quantitative measures: Pearson’s correlation coefficient (r), coefficient of determination (r²), root mean squared error (RMSE), and mean absolute error (MAE).

Pearson’s correlation coefficient measures the strength and direction of the linear relationship between two variables. The coefficient of determination is the square of this coefficient, providing a quantitative measure of how well one variable explains another. This metric ranges from 0 to 1, where 1 represents a perfect fit and values near 0 indicate little to no association. In this context, r² assesses how closely the simulated individual characteristics, aggregated by census tract, align with the actual census tract demographic data. Since the IPF approach is designed to fit individual-level data to the marginal totals in census tract data, it is unsurprising that the values of r and r² for gender, race, age, education, and income are very high for both surveys. Because vaccine uptake is typically unavailable at the census tract level and cannot be directly incorporated into the IPF, our approach “carries over” individual vaccine status along with their attitudes, beliefs, and perceptions during the sampling and replication stage (see Figure 2).

We find that combining IPF with either of the public health surveys allows us to initialize agents with COVID-19 vaccination status in a way that reasonably reflects the real population. The Pearson correlation coefficient and the coefficient of determination for vaccine uptake evaluating the populations generated from both surveys are moderately high (see Table 3). This is visually depicted in Figure 3, where each scatterplot point represents one of the 1,592 Virginia census tracts, with the x-axis showing the observed percentage of vaccine uptake and the y-axis showing the percentage within the synthesized population. In general, within the simulated population using the HPS with IPF, census tracts with higher real vaccination rates also show higher proportions of vaccinated synthetic individuals, with a moderate positive correlation (r = 0.75, r^2^ = 0.56, Figure 3A). A similar pattern emerges in the simulated population from the local survey (r = 0.72, r^2^ = 0.51, Figure 3B). Additionally, when comparing the count of simulated vaccinated individuals in each census tract to the actual count, we find a stronger positive correlation for the HPS dataset (r = 0.91, r^2^ = 0.83, Figure 3C) and the local survey dataset (r = 0.88, r^2^ = 0.77, Figure 3D). However, this is largely a reflection of how well the IPF simulates the total population in each census tract, as larger populations naturally lead to more vaccinated individuals.

**Figure 3.**
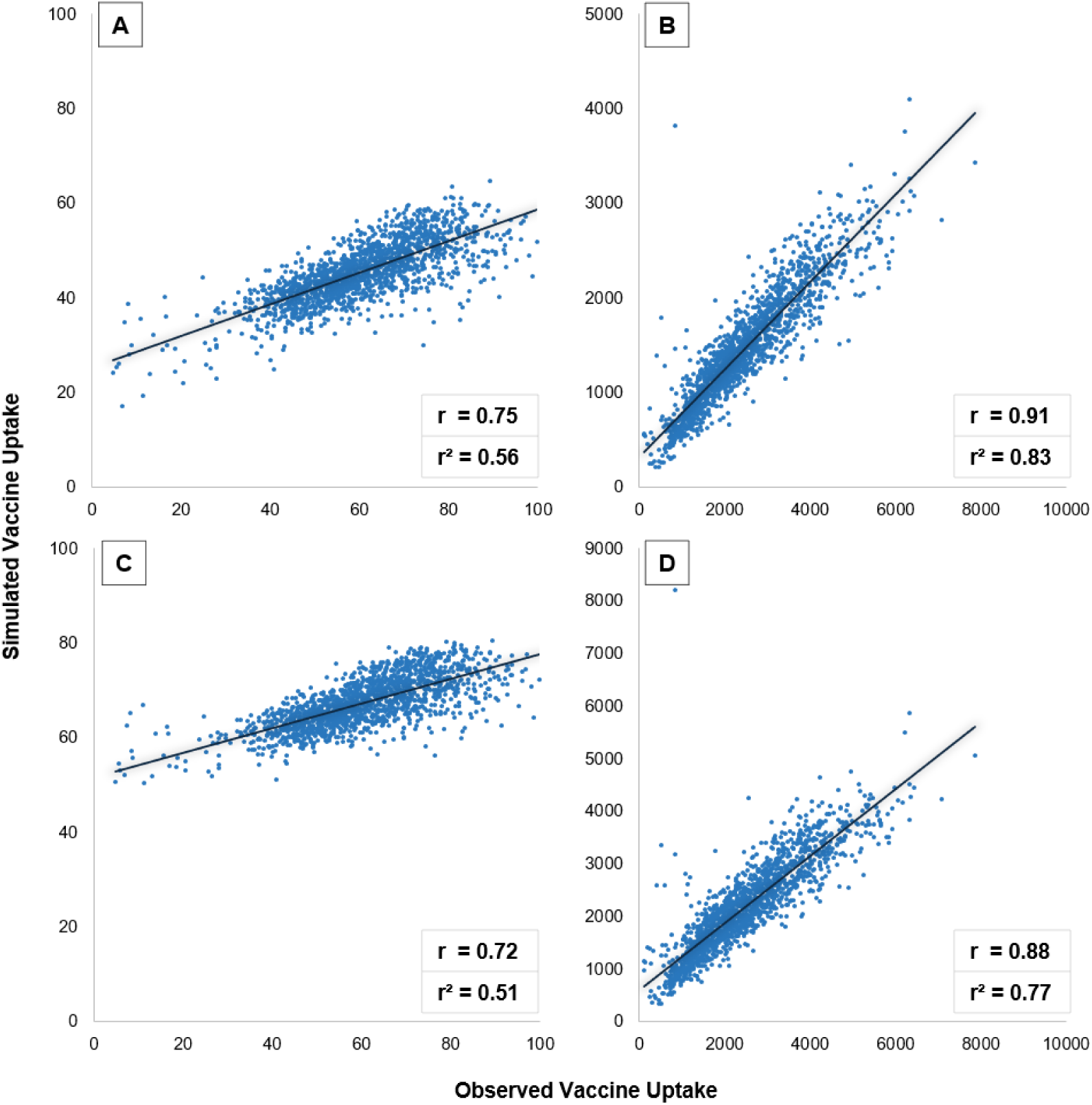
Scatterplots comparing observed and simulated vaccine uptake within Virginia Census Tracts: A) percentage from the HPS, B) count from the HPS, C) percentage from the local survey, D) count from the local survey.

**Table 3.**
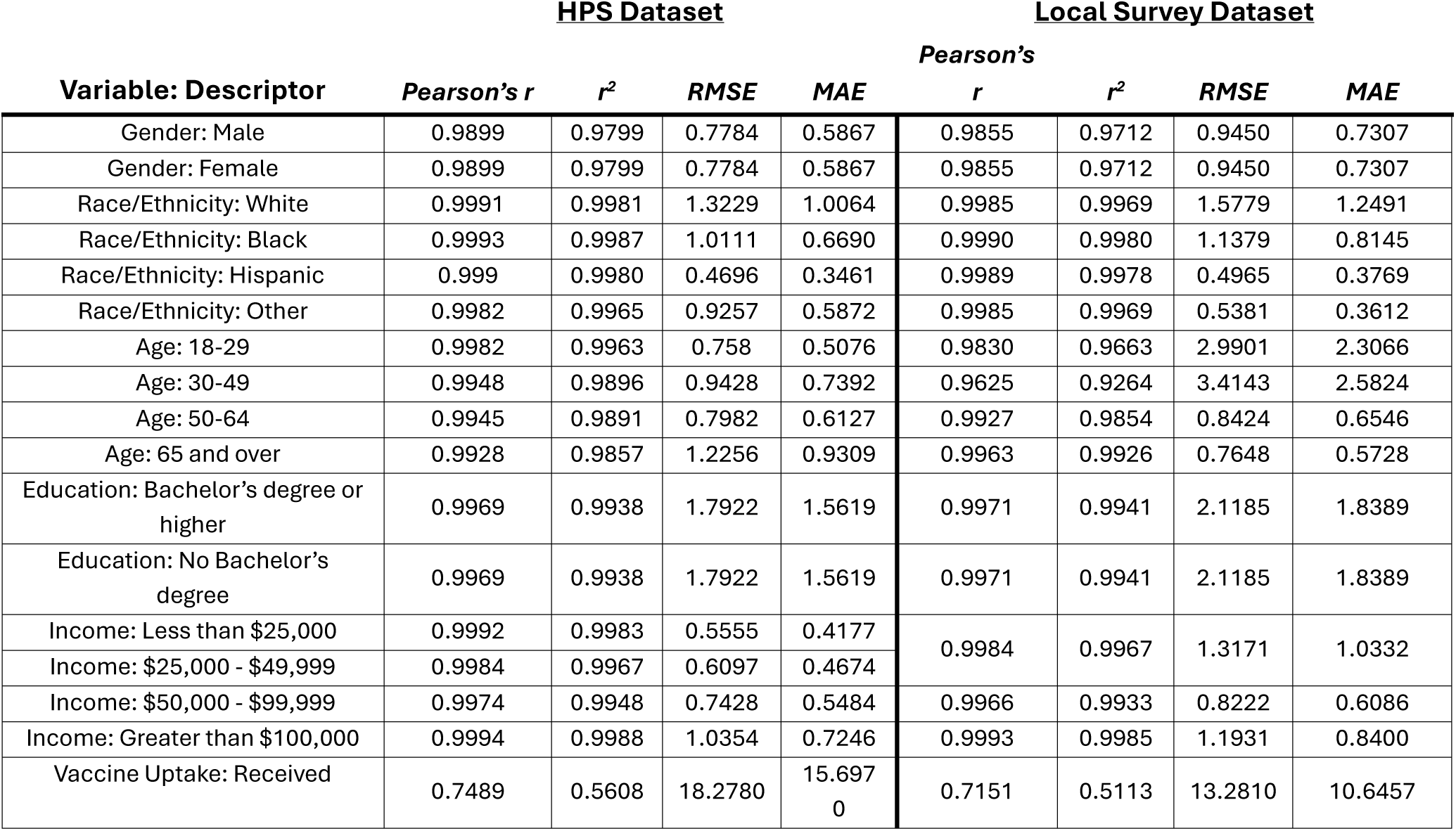
Evaluation metrics comparing observed and simulated percentages of demographic and vaccine status variables across Virginia Census Tracts.

RMSE, measured in the same units as the original data, indicates how closely a simulated population matches the actual census tract (CT) data, with lower values reflecting a better fit and higher values signaling greater discrepancies. As expected, the RMSE values are low for gender, race, age, education, and income. However, the RMSE for the observed and simulated percentage vaccination rates across CTs is 18.28 for the HPS dataset and 13.28 for the local survey dataset. These values suggest that, on average, the simulated percentage of vaccinated individuals differs from the actual percentage by 18.28% and 13.28%, indicating a moderate level of inaccuracy.

Similarly, MAE measures the average magnitude of errors between predicted and observed values by averaging the absolute differences, without considering their direction. Unlike RMSE, MAE does not square the errors, making it less sensitive to large deviations and more robust to outliers. MAE values are consistently low for gender, race, age, education, and income variables in both synthetic populations, as IPF effectively fitted these variables to the CT data. For vaccine uptake percentages, MAE values for the synthesized populations are 15.70 for the HPS dataset and 10.65 for the local dataset, which are comparable to the RMSE values. This indicates that the simulated vaccine uptake percentages differ from actual values by 15.70% and 10.65%, respectively, and the similarity between MAE and RMSE values suggests that large deviations do not disproportionately impact the average error. Overall, both RMSE and MAE suggest that the simulated vaccine uptake percentages from our synthetic population generation approach are reasonably close to the observed values across Virginia census tracts. Furthermore, the RMSE and the MAE are smaller for the local survey dataset, possibly since the dataset is representative of Virginia rather than a national dataset. In general, the synthetic census tracts tend to have a smaller proportion of vaccinated individuals than compared to the real population. This may be attributed to the fact that the validation dataset captures vaccination age 12+ and we simulate agents age 18+.

In contrast, the null model shows significantly poorer performance in initializing agents realistically with vaccine decisions, as evidenced by a Pearson correlation coefficient of 0.298 and a coefficient of determination of 0.089. These low values indicate a weak relationship between the simulated and observed vaccine uptake percentages. Additionally, the null model’s RMSE of 30.357 and MAE of 24.008 are considerably higher compared to our proposed approach. These error metrics suggest greater deviations between the simulated and actual vaccine uptake data in CTs, demonstrating that the null model fails to accurately reflect the real distribution of vaccine uptake. This comparison highlights the limitations of the null model in capturing vaccination behaviors when initializing an agent population and emphasizes the improved performance of our synthetic population generation approach using public health surveys.

Our approach effectively preserves the real-world statistical relationship between sociodemographic variables and vaccine uptake. This is demonstrated by comparing logistic regression coefficients that explain the relationship between these variables and vaccine uptake across the original survey populations, the synthetic population generated with our approach, and the null model. As shown in Table 4, in the HPS dataset, real individuals who are white, male, or low-income (less than $25,000) have lower vaccination rates (β = -0.2277, -0.0595, -0.5776, respectively), while those who are 65 or older and hold a bachelor’s degree or higher (β = 1.1670, 1.2429, respectively) are more likely to be vaccinated. The direction and the relative strength of these associations are also reflected in the synthetic population generated using the HPS dataset. In contrast, the null model fails to capture these underlying statistical relationships. For example, in the synthetic population created by the null model, agents aged 65+ are less likely to be vaccinated (β = -0.11). Similarly, while a bachelor’s degree or higher is strongly associated with increased vaccine uptake (β = 1.24) in the HPS data, the null model results in a weaker association between education attainment and vaccine uptake (β = 0.24).

**Table 4.** Coefficients from logistic regression models describing the relationship between demographic variables and vaccine uptake from the HPS, along with the synthetic population and null model generated from the HPS. Significant coefficients are indicated with an asterisk (*) at the 90% confidence level (p-value < 0.10).

| Variable: Descriptor | <u>HPS Data: $R^2 = 0.08$</u><br>Coefficient $\beta$ | <u>Synthetic Population: $R^2 = 0.11$</u><br>Coefficient $\beta$ | <u>Null Model: $R^2 = 0.01$</u><br>Coefficient $\beta$ |
| --- | --- | --- | --- |
| Intercept | 1.8615* | - 0.3245* | - 0.2365* |
| Gender: Male | - 0.0595* | - 0.1872* | 0.0025 |
| Race/Ethnicity: White | - 0.2277* | - 0.2758* | - 0.2761* |
| Age: 65 and over | 1.1670* | 1.4072* | - 0.1128* |
| Education: Bachelors or Higher | 1.2429* | 1.3435* | 0.2357* |
| Income: \$25,000 or less | - 0.5776* | - 0.8446* | - 0.3071* |

Similar results are presented with the synthetic population generated from the local survey dataset and the corresponding population from the null model (Table 5). In the local survey, individuals who are either aged 65 and older, have a bachelor’s degree or higher, or with high income (greater than $100,000) are more likely to be vaccinated (β = 1.0524, 0.6248, 0.4058, respectively), while individuals who are white are less likely to be vaccinated (β = -0.1511). These associations are reflected in the synthetic population generated using our approach. However, the null model does not capture the strong positive relationship between age 65+ and vaccine uptake observed in the local survey dataset (β = 1.0524), with the coefficient becoming negative and close to zero (β = - 0.0841).

**Table 5.** Coefficients from logistic regression models describing the relationship between demographic variables and vaccine uptake from the local survey, the synthetic population generated from the local survey, and the null model generated from the HPS. Significant coefficients are indicated with an asterisk (*) at the 90% confidence level (p-value < 0.10); Gender was not included due to its insignificance (p-value greater than 0.10) in the local survey dataset.

| Variable: Descriptor | <u>Local Data: $R^2 = 0.05$</u><br>Coefficient $\beta$ | <u>Synthetic Population: $R^2 = 0.05$</u><br>Coefficient $\beta$ | <u>Null Model: $R^2 = 0.01$</u><br>Coefficient $\beta$ |
| --- | --- | --- | --- |
| Intercept | 0.2341* | 0.4020* | - 0.4055* |
| Race/Ethnicity: White | - 0.1511* | - 0.3036* | - 0.2964* |
| Age: 65 and over | 1.0524* | 1.1722* | - 0.0841* |
| Education: Bachelors or Higher | 0.6248* | 0.3839* | 0.1410* |
| Income: Greater than \$100,000 | 0.4058* | 0.6669* | 0.3703* |

It is important to note that the logistic regression examples illustrate how the synthetic populations generated with our public health approach are compared to the real populations from the surveys. Variables for comparison were selected based on their significance in the original surveys, while gender was excluded from the local survey dataset comparison due to its lack of significance at a 90% confidence level. While the strength and direction of the association between sociodemographic variables and vaccine uptake were preserved in the synthetic populations from both the HPS and local survey datasets, the coefficient of determination (R^2^) for the logistic regression also remained relatively consistent, indicating a similar fit between sociodemographic variables and vaccine uptake. Specifically, the R^2^ was 0.08 for the HPS dataset and 0.11 for the corresponding synthetic population, while it was 0.05 for the local survey dataset and the corresponding synthetic population. In contrast, the null models produced a much lower R^2^ of 0.01.

Figure 4 illustrates the spatial heterogeneity of vaccination uptake across the 1,592 census tracts for which data was available for both real and synthetic populations. In these maps, a “High-High Cluster” (light pink) indicates that census tracts have high vaccine uptake and are surround by counties with similarly high vaccine uptake. In contrast, a “Low-Low Cluster” (light blue) represents census tracts with low vaccine uptake and are surrounded by counties also with low vaccine uptake. Outlier census tracts are identified as “High-Low Outliers” (bright red), where census tracts with high uptake are surrounded by those with low uptake, or “Low-High Outliers” (bright blue), where census tracts with low uptake are surrounded by those with high uptake. Census tracts without a significant relationship to their neighbors are shown in light yellow, while those with no population or vaccine data are in grey.

**Figure 4.**
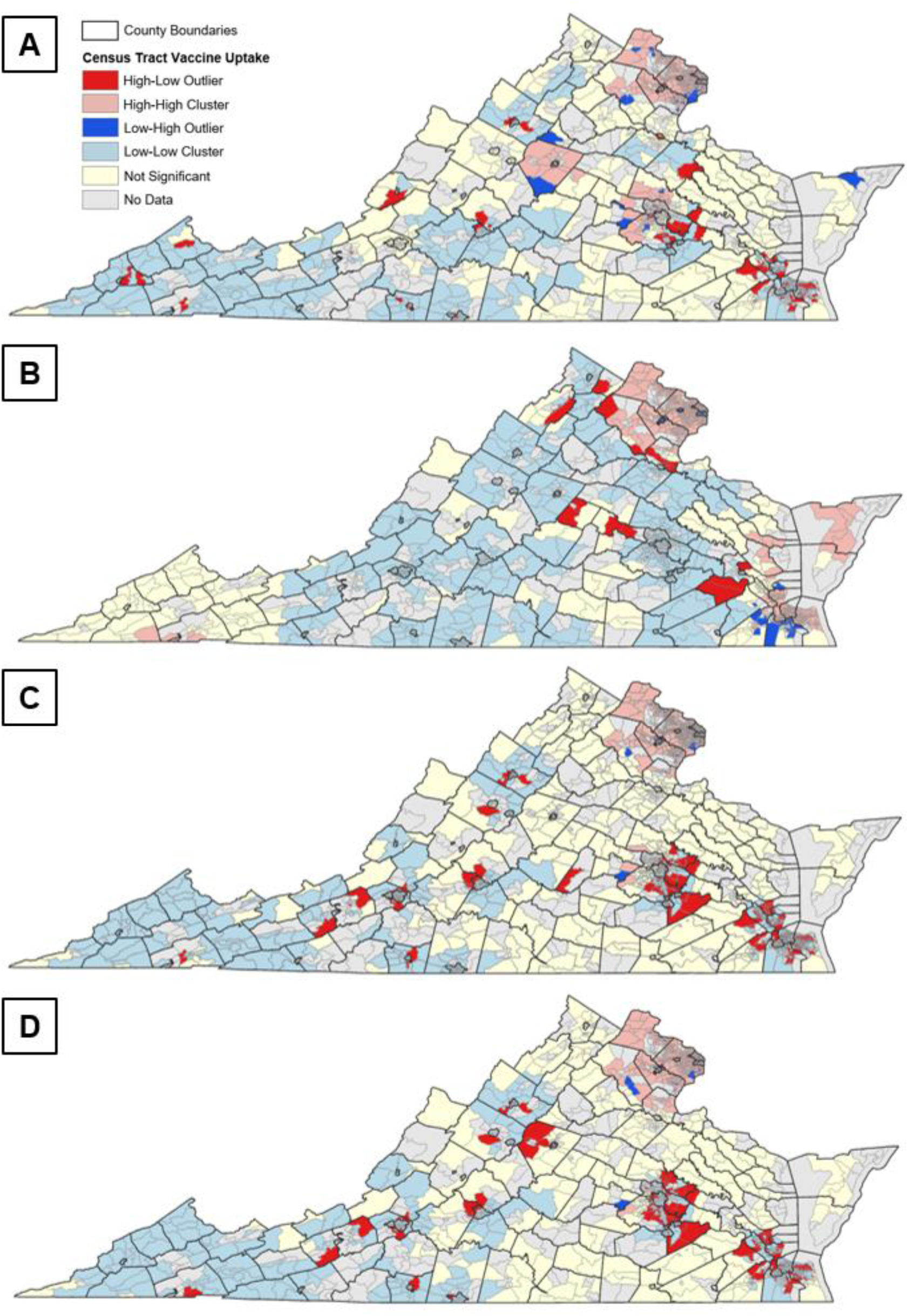
Clusters and outliers of vaccination uptake in Virginia Census Tracts: A) observed, B) null model, and C) synthesized population with HPS data, and D) synthesized population with local survey data.

The observed vaccine uptake by December 2021 is mapped in Figure 4A. Generally, census tracts in the western part of Virginia show relatively low vaccine uptake. Clusters of tracts with high vaccine uptake are found in Northern Virginia, including Fairfax, Prince William, Loudoun, and Arlington Counties. Other high uptake clusters appear in the central part of the state, such as Albemarle County, which surrounds Charlottesville, and Hanover County, particularly in census tracts west of Richmond. The rest of Virginia exhibits mixed uptake rates, leading to the formation of outliers. These outliers are scattered throughout the state, with many High-Low outliers concentrated in larger areas, such as southeast of the Richmond metropolitan area, around Hampton Roads, and in smaller regions near major cities like Harrisonburg and Forest.

Generally, the population generated using the null model approach captures the spatial heterogeneity of COVID-19 vaccine uptake since the marginal totals of the synthetic population vaccination are imposed to match the real county-level data (Figure 4B). However, given that only county-level data is publicly available, there is less within-county variation. For instance, the null model accurately detects the high vaccine uptake cluster in Northern Virginia but inaccurately suggests similar clusters in the southeastern region and census tracts along the Chesapeake Bay. Additionally, the null model overlooks the low vaccine uptake clusters in southwestern Virginia. It also fails to replicate the general outlier patterns observed in real vaccine uptake (Figure 4A), and specifically misclassifies High-Low outliers in southeastern Virginia as Low-High outliers. This indicates that imposing vaccine decisions during the initialization of agent populations does not adequately preserve the spatial distribution of protective behaviors.

The synthetic populations generated using the HPS survey (Figure 4C) and the local survey (Figure 4D) generally align better with observed vaccine rates compared to the null model. They effectively capture the high vaccination cluster in Northern Virginia and the low vaccine cluster in the southwest. However, they fall short in replicating the larger high vaccination clusters in central Virginia near Charlottesville and Richmond. Despite this, our approach excels in preserving both broad regional patterns and location-specific outliers. For example, the High-Low outliers in the Hampton Roads and Richmond metropolitan areas, as well as certain census tracts near Harrisonburg and Forest, are accurately reflected in the synthetic populations. Notably, our method also identifies the sole Low-High outlier census tract west of Richmond, an area primarily characterized by High-Low outliers. These findings highlight the effectiveness of our approach in capturing the spatial heterogeneity of protective behaviors.

With our approach, all variables from the public health survey are incorporated into the agent population, enabling us to generate synthetic populations with not only initial uptake of protective behaviors like vaccination but also realistic attitudes, beliefs, and perceptions. This allows for a better understanding of the spatial patterns of these characteristics within a population. Figure 5 illustrates the vaccination attitudes, beliefs, and perceptions of the synthetic population generated from the HPS survey. For example, in western Virginia, clusters of individuals exhibit vaccine hesitancy due to reasons such as lack of doctor recommendation (Figure 5B), distrust in the vaccine (Figure 5D), or concerns about side effects (Figure 5F). In contrast, Northern Virginia shows a low clustering of individuals planning to wait to see if the vaccine is safe (Figure 5A) or doubting its efficacy (Figure 5H). However, there is a high concentration of individuals who do not perceive COVID-19 as a significant threat (Figure 5C) or do not feel the need for the vaccine (Figure 5G). Concerns about vaccine cost are prevalent in eastern Northern Virginia and extend slightly south, as well as in the southeast around the Hampton Roads region (Figure 5E). Specific census tracts with individuals that believe it is hard for them to get a vaccine are shown in Figure 5I. This approach facilitates the integration of behavioral theories, such as the Health Belief Model (HBM), into ABMs by illustrating how individual attitudes, beliefs, and perceptions affect vaccine uptake and spatial distribution. This capability ultimately supports the development of ABMs of infectious disease spread that aim to simulate the underlying processes driving the adoption of protective behaviors over time, providing a realistic initialization of populations with these characteristics.

**Figure 5.**
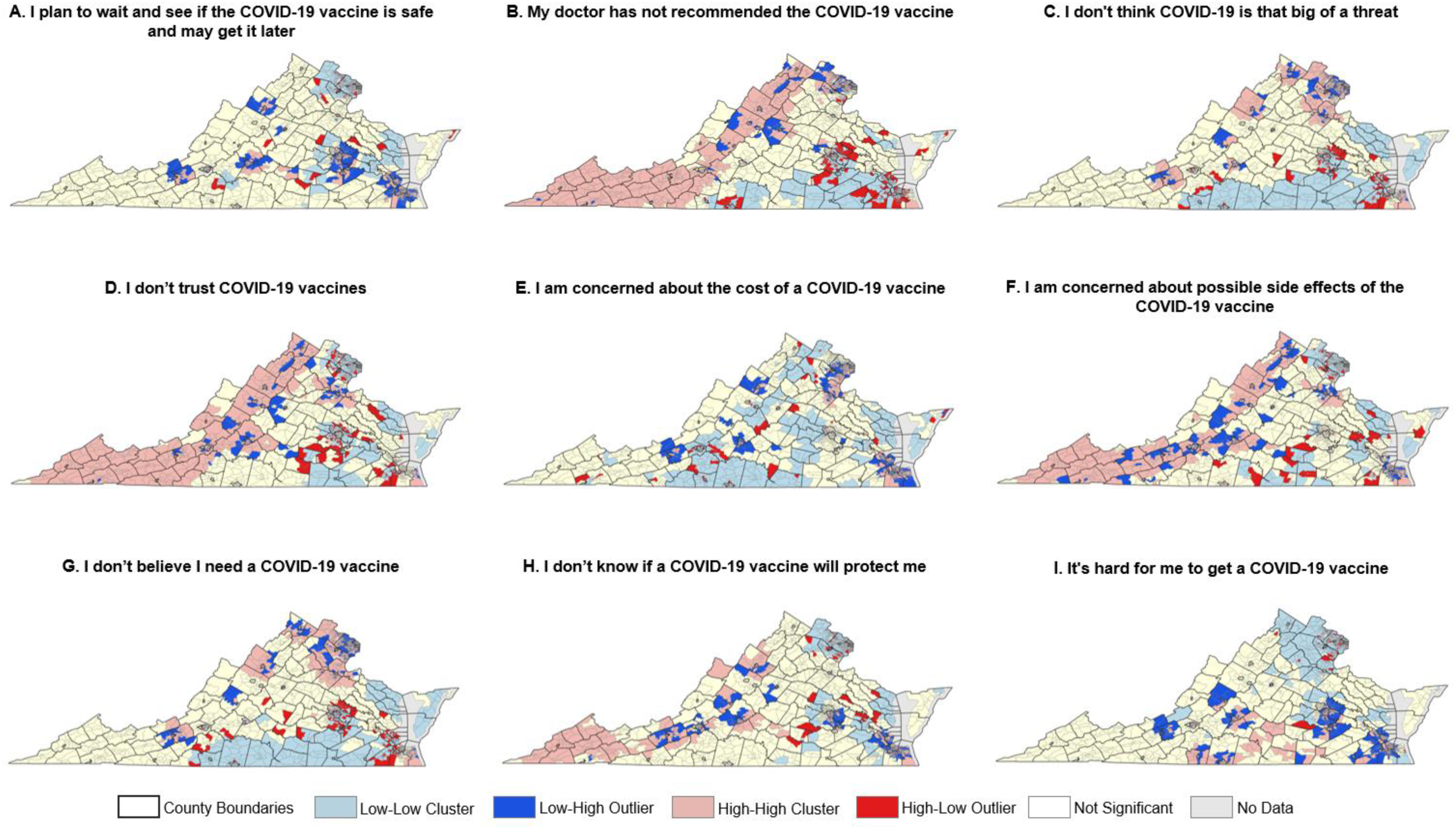
Cluster and outliers of vaccination attitudes, beliefs, and perceptions within Virginia Census Tracts for the synthetic population generated from the HPS.

## 5. Discussion and Conclusion

In this study, we investigate the potential to expand synthetic population generation approaches to initialize an agent population with variables relevant for public health, using COVID-19 vaccine uptake as an example. Our results show that such an approach has potential to support disease simulations requiring realistic parameterization of agents with these variables. This method enables researchers to quickly initialize a synthetic population where the true statistical relationships between demographic characteristics and public health variables are preserved. Furthermore, the approach captures the spatial heterogeneity of such protective behaviors at finer scales than typically available in spatial data. While protective behaviors such as vaccination, masking, and social distancing can sometimes be found at county or state level, similar data capturing attitudes, beliefs and perceptions that can be simulated using this approach are often not available in spatial data format at all. The synthetic population that was generated using the national HPS that is publicly available was comparable to the synthetic population generated using the local survey data, demonstrating the flexibility of the approach to be implemented using a variety of public health surveys.

It is important to note that researchers who have access to fine-grained spatial data capturing health behavior variables (e.g. vaccine uptake at the CT level) could incorporate that data directly into the IPF approach to more accurately capture the statistical and spatial patterns health behaviors. However, this would be limited to the study area and time for which the data is available. For example, our validation dataset captures vaccine uptake at the CT level for Virginia by December 2021, meaning it could be used in the IPF as another category for which the marginal totals are known. However, this would limit the transferability of the approach to other study areas and points in time. Therefore, we demonstrate how the approach could be implemented using only publicly available longitudinal data such as the HPS, making it straightforward for researchers to generate a synthetic population with these variables anywhere in the country and for multiple points in time.

The quality of the synthetic population is limited by the quality of the census tract and individual level survey data. For example, it does not appear that the HPS data is nationally representative and was largely biased towards vaccinated individuals. Therefore, we were able to improve our results slightly using this dataset by adjusting the representation of vaccinated individuals in the sample from 91% (r^2^ = 0.499) to 50% (r^2^ = 0.56), but more research is needed to investigate effects of bias on this approach. Furthermore, there are other factors that are likely affecting the individual’s decision to get vaccinated (e.g. policy interventions, social norms) that can’t be directly incorporated into the synthetic population generation approach. Thus, it may be more effective to synthesize a population with vaccine intention, rather than vaccine uptake itself, where such data is available (e.g. using the Understanding Coronavirus in America longitudinal survey).

In conclusion, this study demonstrates the potential of using public health surveys to enhance the generation of synthetic populations for spatial agent-based models (ABMs) by incorporating protective behaviors and attitudes. Such an approach strengthens the potential for ABM in public health research and policy planning. We show that the synthetic populations generated using this approach reflect the real-world statistical relationships between demographic groups and vaccine uptake and attitudes. This level of detail is essential for simulating potential health disparities across different demographic groups, enabling the exploration of more targeted and effective public health strategies. Initializing an ABM with realistic vaccine uptake and attitudes is crucial for those that aim to forecast outbreaks in study areas where uptake and attitudes vary regionally or that aim to simulate realistic social processes driving vaccine uptake decisions. By capturing the spatial heterogeneity of these behaviors and attitudes at finer scales than spatial data typically allows, our approach supports models that aim to simulate how local responses to interventions might unfold with greater accuracy and how these responses lead to spatially heterogeneous health outcomes. Ultimately, this approach enhances the predictive power and realism of disease simulations, providing critical insights into how interventions might play out in real-world settings.

To our knowledge, this study marks one of the first attempts to extend synthetic population generation approaches to initialize agents with variables relevant to ABMs of infectious disease spread. Future research is needed to see if this approach can be used to initialize other health behaviors and associated perceptions and attitudes (e.g., tobacco use in populations using the CDC’s National Tobacco Survey). We encourage other researchers with access to more fine-grained spatially aggregated data to validate this approach across various public health domains, to improve the parameterization of more realistic agent populations in data-driven ABMs for public health.

## Acknowledgements

This research was funded by National Science Foundation (Award #230970 and #2109647).

## Data availability statement

The data including the validation dataset and code for generating the synthetic population based on the Household Pulse Survey is available on a GitHub repository at https://github.com/evonhoene/Population-Generation-for-Public-Health-ABMs. The spatially aggregate data from the American Community Survey and the individual-level Household Pulse Survey are publicly available and the links are provided on the GitHub page. De-identified data from the local Virginia survey data is available upon request.

